# Prevalence of SARS-CoV-2 and co-occurrence/co-infection with malaria during the first wave of the pandemic (the Burkina Faso case)

**DOI:** 10.1101/2022.09.20.22280138

**Authors:** Diana López-Farfán, R. Serge Yerbanga, Marina Parres-Mercader, Manuela Torres-Puente, Inmaculada Gómez-Navarro, Do Malick Soufiane Sanou, Adama Franck Yao, Jean Bosco Ouédraogo, Iñaki Comas, Nerea Irigoyen, Elena Gómez-Díaz

## Abstract

Africa accounts for 1.5% of the global coronavirus disease 2019 (COVID-19) cases and 2.7% of deaths, but this low incidence has been partly attributed to the limited testing capacity in most countries. In addition, the population in many African countries is at high risk of infection with endemic infectious diseases such as malaria. Our aim is to determine the prevalence and circulation of SARS-CoV-2 variants, and the frequency of co-infection with the malaria parasite. We conducted serological tests and microscopy examinations on 998 volunteers of different ages and sexes in a random and stratified population sample in Burkina-Faso. In addition, nasopharyngeal samples were taken for RT-qPCR of SARS-COV-2 and for whole viral genome sequencing. Our results show a 3.2% and a 2.5% of SARS-CoV-2 seroprevalence and PCR positivity; and 22% of malaria incidence, over the sampling period, with marked differences linked to age. Importantly, we found 2 cases of confirmed co-infection and 8 cases of suspected co-infection mostly in children. Finally, we report the genome sequences of 13 SARS-CoV-2 isolates circulating in Burkina Faso at the time of analysis, assigned to lineages A.19, A.21, B.1.1.404, B.1.1.118, B.1 and grouped into clades; 19B, 20A and 20B. This is the first population-based study about SARS-CoV-2 and malaria in Burkina Faso during the first wave of the pandemic, providing a relevant estimation of the real prevalence of SARS-CoV-2 and variants circulating in this Sub-Saharan African country. Besides, it highlights the low frequency of co-infection with malaria in African communities.

## 1 Introduction

To date, the SARS-CoV-2 coronavirus, responsible of the coronavirus disease 19 (COVID-19), has caused more than 606 million cases and 6,5 million deaths worldwide, a small proportion of these (9,3 million cases [1.5%] and around 174,407 deaths [2.7%]) have occurred in Africa (data as of September 13th 2022, WHO^1^). Despite their under-funded health systems, cases and deaths reported in Africa appear low compared to other parts of the world. In addition, most of cases cluster in few Northern and Southern countries (i.e. South Africa [4,01M cases], Morocco [1,26M cases] and Tunisia [1,14M cases]), while data in most Western African countries is scarce or absent (i.e. the number of accumulative cases considering all countries in Western Africa is ∼936K). However, these estimations should be considered with caution given the low number of tests conducted in most African countries compared to data available for Europe (one or two orders of magnitude higher). A more proper effort to estimate the real incidence is needed to make conclusions about the extent of this pandemic in the African continent.

To prevent the emergence and spread of new variants across the world, sequencing of SARS-CoV-2 lineages circulating in Africa is critical (1). Indeed, the higher rates of COVID-19 testing and genomic surveillance in the south of the continent have permitted the early identification of several variants of concern (VOC) such as Beta (B.1.351) and Omicron (B.1.1.529); and different variants of interest (VOI) such as C.1.1 (2,3). To date, most of the deposited sequences from Africa in the GISAID database come from the South African region (47%), while sequences from Western African countries only account for 17.6% (GISAID^2^ as of 31 May 2022) (4). Such an uneven African genomic surveillance holds a weakness in our global surveillance system and raises the concern of not leaving Africa behind in the global pandemic response (1).

Besides the direct effects of COVID-19 in terms of cases and deaths, the pandemic has shown to have important collateral effects on several other infectious diseases such as HIV/AIDS, tuberculosis and malaria (5). According to the World Health Organization (WHO), half of all deaths in Africa are caused by these three infectious diseases (∼2.4 million people/year) compared to only 2% in Europe. The Sub-Saharan Africa is one of the most affected regions with significant drops in notifications of all three diseases. According to the latest WHO World malaria report^3^, 15 malaria endemic countries reported more than 20% reduction in malaria testing and treatment between April and June of 2020^3^.

Malaria is a life-threatening disease caused by unicellular eukaryotic parasites of the genus Plasmodium that are transmitted to humans through the bites of infected female Anopheles mosquitoes. Sub-Saharan Africa carries a disproportionate portion of the global malaria burden. In 2020, the region was home to 95% of malaria cases and deaths. Children below 5 years are the most vulnerable group affected by malaria; accounting for 80% of all malaria deaths in the region^3^. At the beginning of the COVID-19 pandemic, studies projected that malaria deaths could double in 2020 due to health service disruptions (6). However, many countries took urgent actions to avoid the worst projections. In 2020, there were an estimated 241 million cases of malaria and 627,000 deaths worldwide, this is 14 million more cases and 69,000 more deaths than in 2019. Two-thirds of this increase have been associated to disruption in malaria prevention campaigns, diagnosis and treatment during the first wave of the pandemic^3^. Burkina Faso is a Western African country that confirmed its first community transmission cases of SARS-CoV-2 on March 15, 2020. To date, this country has reported 21,128 total cases which is a very low incidence compared with the 4M of total cases reported from South Africa, for example. Whether this rate is representative of the real epidemiological situation of the country, or if it is due to insufficient testing, remains and it will probably remain unanswered. On the other hand, malaria that is endemic across the country, is a major health issue with a seasonal peak from June to October. During the COVID-19 pandemic, malaria case incidence (cases per 1000 population at risk) increased from 366.1 in 2019 to 389.9 in 2020^4^. The high malaria incidence in this country and the increase in prevalence detected during the first wave of the COVID-19 pandemic raised the question about how frequent has been SARS-CoV-2 and malaria co-infection.

In order to address this, 998 volunteers from different rural and urban areas in the south of Burkina Faso were screened for malaria by microscopy, and for SARS-CoV-2 using rapid diagnostic serology tests and RT-qPCR. In addition, PCR positive samples were whole-genome sequenced and analyzed through comparative genomics and phylogenetics. We found a 3.2% seroprevalence and 2.5% of PCR positivity for SARS-CoV-2 over the studied period. Although age groups mostly affected by each disease do not overlap, 10 cases of SARS-CoV-2/malaria co-infection in younger age groups were found. Finally, we reported 13 SARS-CoV-2 whole genomes circulating in Burkina Faso at the time of sampling. Viral genomes clustered into 3 early clades (i.e. 19B, 20A and 20B) and lineages found were A.19, A.21, B.1.1.404, B.1.1.118 and B.1.

Altogether, our data shed new light on the incidence of COVID-19 and the different SARS-CoV-2 circulating variants in a Western African country, and its co-occurrence with another endemic infectious disease of utter importance such as malaria.

## 2 Methods

### Ethics

This protocol was carried out by the Institut des Sciences et Techniques de Bobo-Dioulasso and was approved by the Spanish National Research Council (CSIC), the Ethics Committee of Biomedical Research in Andalusia (CCEIBA), and the Centre National de la Recherche Scientifique et Technologique (CNRST) of Burkina-Faso. The protocol is in conformity with the declaration of Helsinki and all international regulations about the ethical principles in biomedical research involving human subjects. SARS-CoV-2 seropositive and malaria positive individuals were provided with treatment and assisted by medical professionals. During the screening, local authorities, school directors and families of volunteers were informed about the objectives of the study and protocols performed. As the study involved children, a signed declaration of conformity for the participation in the study was obtained from their parents and/or legal representatives.

### Study design

The study was conducted in 11 villages from different rural and urban areas in the south of Burkina Faso (**Supplementary Figure 1**). From August to November 2020, a total of 998 asymptomatic volunteers were enrolled. Demographic and clinical information collected included age, sex and temperature. For each participating subject, two drops of blood were obtained by capillary puncture of the fingertip. This blood was used for malaria detection by microscopy, and for COVID-19 serology testing using a point of care rapid test (INgezim COVID-19 CROM Easy). When there was consent, nasopharyngeal samples were also collected in AVL buffer (Qiagen) for molecular testing of SARS-CoV-2 using RT-qPCR.

### SARS-CoV-2 diagnosis

Detection of anti-SARS-CoV-2 antibodies was performed on volunteers’ capillary blood using the INgezim COVID-19 CROM Easy kit (Gold Standard Diagnostics), following manufacturer’s instructions. This serology-based finger-prick test is based on a dual-recognition immunochromatographic assay that determines the presence of total antibodies (IgG, IgA, and IgM) specific to SARS-CoV-2 in a single blood sample by using the nucleoprotein (N protein) as antigen. The manufacturer reported specificity of 99.3% and sensitivity of 94.5%. No cross-reactivity with other coronaviruses or agents related to human respiratory illnesses have been reported. In this study, we estimated seroprevalence as the proportion of individuals who had a positive result in the point-of-care test after 10 minutes.

Nasopharyngeal samples were taken using a sterile swab. The swab was immediately placed in a pre-filled tube containing 0.5 mL of inactivating lysis buffer (AVL, Qiagen). Samples were stored at 4ºC for 24h and at -20ºC for prolonged storage for subsequent analysis. Molecular detection of SARS-CoV-2 by RT-qPCR on seropositive volunteers was carried out in the Institut de Recherche en Sciences de la Santé/ Direction Régionale de l’Ouest (IRSS DRO) of Burkina Faso using the FastPlexTriplex SARS-CoV-2 detection kit (gene target identified: ORF1ab, N and the human RNase P as control) (PreciGenome, LLC).

Swab samples of seronegative volunteers were stored at -80ºC and processed in the Institute of Parasitology and Biomedicine Lopez-Neyra (IPBLN) as follows: RNA was extracted from 140 µL of pooled swab samples using the QIAamp viral RNA extraction kit (Qiagen). A pool size of five samples was used in the villages with seropositive subjects (Bobo-Dioulasso and Dandé), whereas samples from villages with negative seroprevalence were processed in pools of 10 samples. RT-qPCR analysis was performed using the one step Direct SARS-CoV-2 Realtime PCR Kit (Vircell S.L) following the manufacturer’s instructions. Targets identified with this kit are N, E and hRNase P genes. RT-qPCR was performed in 20 µL final volume reactions (5 µL of RNA sample) using a CFX96 Real-Time PCR Detection System (Bio-Rad, Hercules, CA, USA). Cycle threshold (Ct) values were analyzed using the BIORAD CFX manager software. Samples of positive pools were extracted individually and retested by RT-qPCR. Samples were considered positive when the N and E target genes had a Ct < 40. Positive and negative controls were included in all experiments.

### Whole Genome Sequencing and Phylogenetic Analysis

RNA from SARS-CoV-2 positive samples with a Ct value ≤35 was sent to sequencing to the Institute of Biomedicine of Valencia (IBV-CSIC). RNA was retrotranscribed into cDNA and SARS-CoV-2 complete genome amplification was performed in two multiplex PCR, accordingly to openly available protocol developed by the ARTIC network^5^ using the V4.1 multiplex primers scheme (artic-network n.d.). Two resulting amplicon pools were combined and used for library preparation. Genomic libraries were constructed with the Nextera DNA Flex Sample Preparation kit (Illumina Inc., San Diego, CA) according to the manufacturer’s protocol with 5 cycles for indexing PCR. Whole genome sequencing was carried out in a MiSeq platform (2×150 cycles paired-end run; Illumina). Sequences obtained went through an open source bioinformatic pipeline based on IVAR^6^ (7). The different steps of the pipeline are as follows: 1) removal of human reads with Kraken (8); 2) filtering of the fastq files using fastp v 0.20.1 (9) (arguments: --cut tail, --cut-window-size, --cut-mean-quality, -max_len1, -max_len2); 3) mapping and variant calling using IVAR v 1.2; and 4) quality control assessment with MultiQC (10). Consensus sequences generated by this pipeline were aligned against the SARS-CoV-2 reference sequence (11) with MAFFT (12). Problematic positions were masked using the mask_alignment.py script from the repository maintained by Rob Lanfear^7^. Lineages and clade nomenclature were assigned using Pangolin (13) and Nexclade online tool (14). Phylogenetic tree was generated with Nextclade v2.5.0^8^, the tree was downloaded in JSON format and visualized using Auspice v2.37.3 online tool^9^ (15,16).

### Malaria diagnosis

Malaria prevalence was estimated in the same population sample of 998 volunteers simultaneously to SARS-CoV-2 detection. Detection of malaria was performed by microscopic examination of blood thick smears that were prepared following standard methods (17). Thick smears were screened for the presence of *Plasmodium falciparum* asexual and gametocyte stages in the blood.

### Statistical analysis

We tested the association between variables with the Wilcoxon Sum Rank tests (for continuous quantitative variables) and the Kruskal-Wallis chi-squared test.

## 3 Results and Discussion

COVID-19 transmission in Africa has been marked by relatively fewer infections, mostly asymptomatic, and lower death rate compared to developed countries. A younger population structure and a variety of socio-ecological factors (i.e. warm weather, low population density and mobility and trained immunity by previous high burden infectious diseases), as well as early public health measures taken by governments (i.e. early lockdown sub-Saharan countries) (18), have been suggested to explain such lower incidence (19). Surprisingly, the pandemic has been more pronounced in a few countries (e.g. South Africa, Morocco and Tunisia) suggesting country-specific drivers of SARS-CoV-2 spread and morbidity, but this could also be largely explained by a low and unequal testing capacity. Reported cases of COVID-19 are dissimilar between countries like South Africa (4,01 million cases and 102,129 deaths), Kenia (338,301 cases and 5,674 deaths) or Ethiopia (493,353 cases and 7,572 deaths). Testing also differs greatly, with 50,000 tests/day in South Africa versus 5,000 tests/day in Kenia or Ethiopia. In Burkina Faso, with a population of 20,903,278, only 21,128 cases and 387 deaths have been reported (as of September 13th 2022, WHO^1^). However, no data about the testing effort in Burkina Faso are available for comparison.

At the beginning of the pandemic, serological surveillance was used as a prospective tool to define the cumulative incidence of COVID-19, particularly in the presence of asymptomatic or mild infections (20). At present, seven countries in Western Africa, four countries in Eastern Africa and four countries in Central Africa have conducted seroprevalence studies that are published or available as preprint (**Supplementary Table 1**). Serological surveys in African countries show highly heterogeneous results with SARS-CoV-2 seroprevalence ranging from 0.9% to 45% during 2020 (**Supplementary Table 1**). To our knowledge, no data have been reported from Burkina Faso for any period.

In this study, we report the results of a population-based and stratified survey involving a total of 998 volunteers that were recruited during a period of 3 months in 2020 (August 22nd to November 19th). The sampling was conducted in 11 villages from different rural and urban areas in the south of Burkina Faso (**Supplementary Figure 1**). 55% of the participants (549 out of 998) were female, and 45% (449 out of 998) were male. The samples were distributed equally into four age groups: (A) 5-12 years (25.2%); (B) 13-20 years (24.9%); (C) 21-40 years (25.2%); and (D) >40 years (24.7%). Importantly, all the participants were asymptomatic. The distribution of positive cases by age and gender classes is detailed in **Table 1**.

**Table 1.** Distribution of positive cases by age classes and gender

| Age group | Sex | non-infected | COVID-19 IgG/A/M | malaria | COVID-19 Ig & malaria | SARS-CoV-2 PCR | SARS-CoV-2 PCR & COVID-19 Ig | SARS-CoV-2 PCR & malaria | Total |
| --- | --- | --- | --- | --- | --- | --- | --- | --- | --- |
| A 5-12 | F | 75 | 0 | 46 | 0 | 0 | 0 | 0 | 121 |
| A 5-12 | M | 67 | 6 | 60 | 3 | 1 | 0 | 1 | 130 |
| B 13-20 | F | 84 | 6 | 34 | 2 | 0 | 0 | 0 | 122 |
| B 13-20 | M | 82 | 1 | 43 | 1 | 3 | 0 | 1 | 127 |
| C 21-40 | F | 150 | 1 | 11 | 0 | 4 | 0 | 0 | 166 |
| C 21-40 | M | 79 | 1 | 4 | 0 | 1 | 0 | 0 | 85 |
| D >40 | F | 116 | 10 | 14 | 2 | 5 | 3 | 0 | 140 |
| D >40 | M | 87 | 7 | 7 | 0 | 10 | 4 | 0 | 107 |
| Sub-total | F | 425 | 17 | 105 | 4 | 9 | 3 | 0 | 549 |
| Sub-total | M | 315 | 15 | 114 | 4 | 15 | 4 | 2 | 449 |
| **Total** |  | **740** | **32** | **219** | **8** | **24** | **7** | **2** | **998** |

A total of 32 individuals out of 998 tested positive for SARS-CoV-2 total antibodies (IgG/A/M), resulting in an overall seroprevalence rate of 3.2% (95% CI 2.1–4.3). However, most of seropositives came from the urban city of Bobo-Dioulasso where local seroprevalence was of 4.96% at that time. A higher seroprevalence in urban areas in Africa has been reported previously (21–24). Apart from urban vs. rural differences, we report significant differences between age groups in SARS-CoV-2 seropositivity (Kruskal-Wallis chi-squared=16.285, df=3, p-value < 0.001). The highest seroprevalence was in the group age of >40 years (6.9% [95% CI 3.7–10.1]), followed by teenagers aged 13-20 years (2.8% [95% CI 0.7–4.9]) and children aged 5–12 years (2.4% [95% CI 0.5–4.3]), whereas the lowest seroprevalence was observed in the group of 21-40 years (0.8% [95% CI 0–1.9]). (**Figure 1A, Table 1**). However, there were no significant differences in SARS-CoV-2 seroprevalence between female and male individuals (Wilcoxon rank sum test W = 15690, p-value = 0.78) (**Supplementary Figure 2**), which agrees with other population-wide seroprevalence surveys in Sub-Saharan Africa (22,25,26). Finally, we also observed that SARS-CoV-2 seroprevalence varied markedly with time with a significant increase in the incidence with a maximum of ∼5.5% in November 2020 (**Supplementary Figure 3**). This increase in infection rate agrees with previously published reports from the African CDC and WHO^1^ that observed a gradually increase in cases since September 2020.

**Figure 1.**
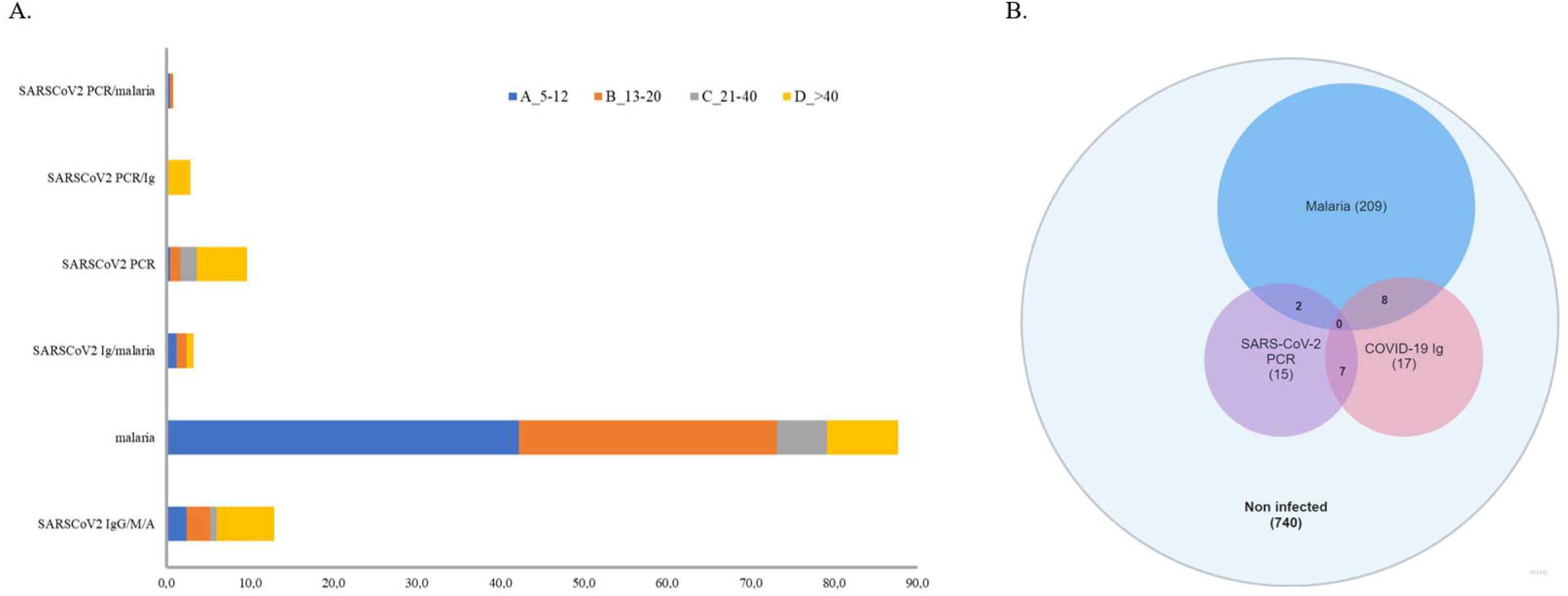
**(A)** Prevalence of SARS-CoV-2, malaria and co-infection by age group. **(B)** Venn diagram representation of positive cases.

Furthermore, seroprevalence estimates for SARS-CoV-2 obtained in this study are also comparable with results reported in other African countries during 2020 (although sample period and the testing effort might be different) (**Supplementary Table 1**). For example, in a serosurvey of asymptomatic people conducted in May 2020 in Addis Ababa (Ethiopia), 3.0% tested positive for SARS-CoV-2 IgG (27). Similarly, a seroprevalence study of 3,098 Kenyan blood donors sampled between April-June 2020 reported a national seroprevalence of 4.3%, with some urban regions reaching around 8% (e.g. Nairobi and Mombasa) (21). Another study in Ethiopia during July 2020 in 14 urban areas found a seroprevalence of 3.5% from 16,932 samples (22).

The 32 SARS-CoV-2 seropositive individuals were subjected to RT-qPCR. SARS-CoV-2 active infection was confirmed in 7 samples (seropositive/PCR positive). Due to the low sensitivity of rapid antibody tests to detect early infections, all the seronegative samples were also analyzed by RT-qPCR. Swab samples were first processed in pools, and then, positive pools were processed individually. Pooled testing is a useful approach to reduce time, cost and increase testing capacity in areas with low prevalence of COVID-19. This PCR pooling works with maximum efficiency with a prevalence <3% and a pool size <10 (28,29). First, we tested the sensitivity of our ‘pool-test’ in groups of five (one positive sample diluted in four negative ones) and ten (one positive sample diluted in nine negative ones) samples (**Supplementary Table 2**). With these results, we decided to group samples in pools of five in areas with seropositive subjects (i.e. Bobo-Dioulasso and Dandé); whereas samples from areas with zero seroprevalence, pools of 10 swabs were processed instead. RNA was extracted from pooled swab samples and RT-PCR was performed. Then, samples in positive pools were extracted and tested individually. With this procedure, we found 17 additional SARS-CoV-2 positive samples by RT-qPCR, most of them from the urban locality of Bobo-Dioulasso (**Table 1, Figure 1B)**. In concordance with serological tests, there were not significant differences in SARS-CoV-2 PCR positivity between females and males (Wilcoxon rank sum test W = 125348, p-value = 0.08114) (**Supplementary Figure 2**). However, there were significant differences between age groups (Kruskal-Wallis chi-squared = 20.155, df = 3, p-value < 0.001) with the highest prevalence in the age group of >40 years decreasing with age. The lowest prevalence was observed in children between 5–12 years (**Figure 1A, Table 1**).

19 out of 24 SARS-CoV-2 PCR positive samples had a Ct value ≤ 35 and they were processed for whole-genome sequencing. After genome assembly and quality control processes, good quality genomes were obtained from 13 samples. To assign the sequences to lineages and clades, we used the dynamic lineage classification method called Phylogenetic Assignment of Named Global Outbreak Lineages (PANGOLIN) (13) and the sequence analysis web tool, Nextclade (14). Phylogenetic analysis showed that our SARS-CoV-2 genomes clustered into three clades: 19B, 20A and 20B (**Table 2, Figure 2**). Most of the samples belong to clade 19B, this one along with 19A were the clades that emerged in Wuhan in late 2019 and dominated the early outbreak. Clade 20A emerged from 19A and dominated the European outbreak in March 2020, and 20B is a sub-clade of 20A (30). Lineages found were A.19, A.21, B.1.1.404, B.1.1.118 and B.1. The distribution and description of these lineages^10^ are summarized in **Supplementary Table 3**. Lineages A.19, A.21 and B.1.1.404 were common in Burkina Faso at the time of analysis, and importantly, were described there for the first time^10^. However, lineages B.1.1.118 and B.1 were not common in Burkina Faso nor even in Africa at that time^10^. As these lineages presented a worldwide distribution, they probably correspond to imported lineages. Indeed, B.1 was reported in Morocco, Saudi Arabia, Spain, France and Brazil on February 2020^11^. A genomic study of SARS-CoV-2 conducted in Ghana with samples from March 12 to April 1 2020 and the end of May 2020, reports the same clades that we found circulating in Burkina Faso from August to November 2020, moreover they also report the linage B.1 (31). Although little divergence was observed (**Figure 2**) as sequences comes from early days in the pandemic, we detected a common spike mutation in most of the Burkina Faso genomes, D614G mutation (**Table 2**). A SARS-CoV-2 variant carrying this mutation became dominant during the early days of the pandemic at global level and had a presumed fitness advantage (32). The D614G mutation was also dominant in Guinea genomes (31) and north-middle African genomes (33).

**Table 2.**
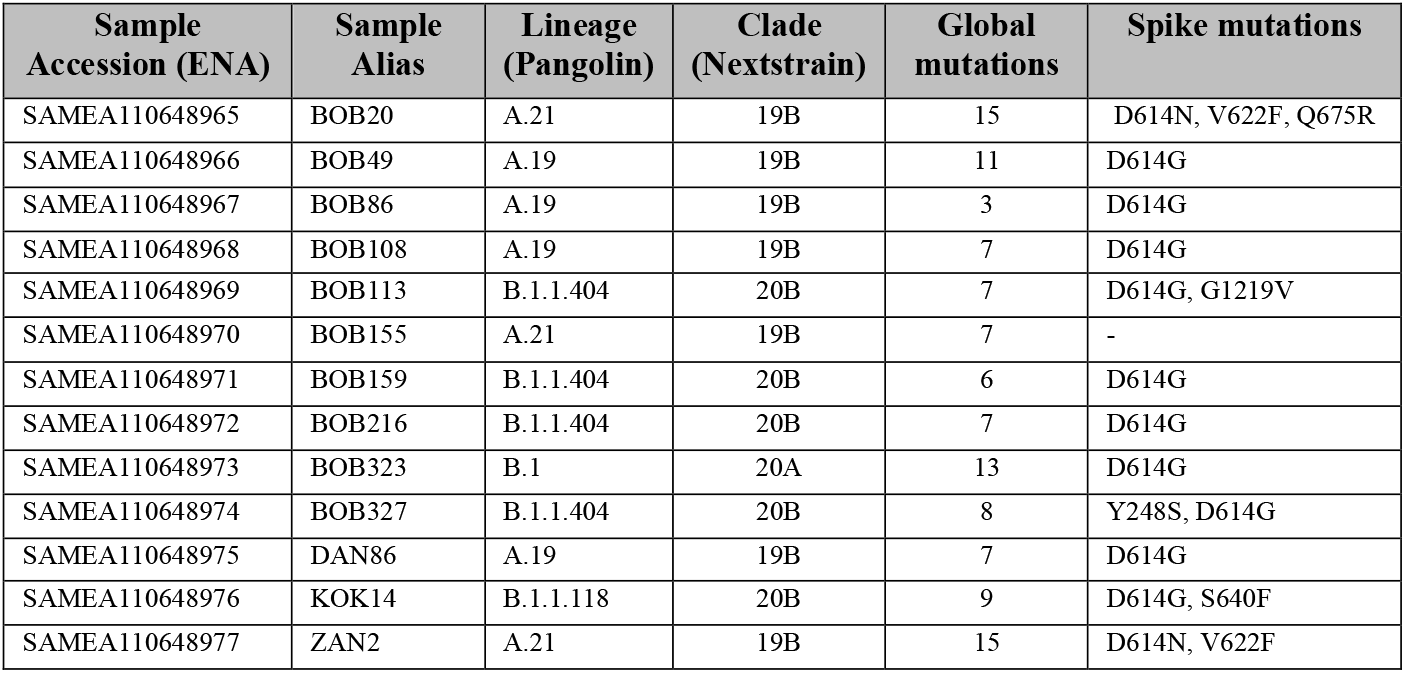
Circulating lineages and clades of SARS-CoV-2 in Burkina Faso from August to November 2020. Mutations (aminoacid substitutions) relative to the reference sequence Wuhan-Hu-1/2019 (MN908947) are showed.

**Figure 2.**
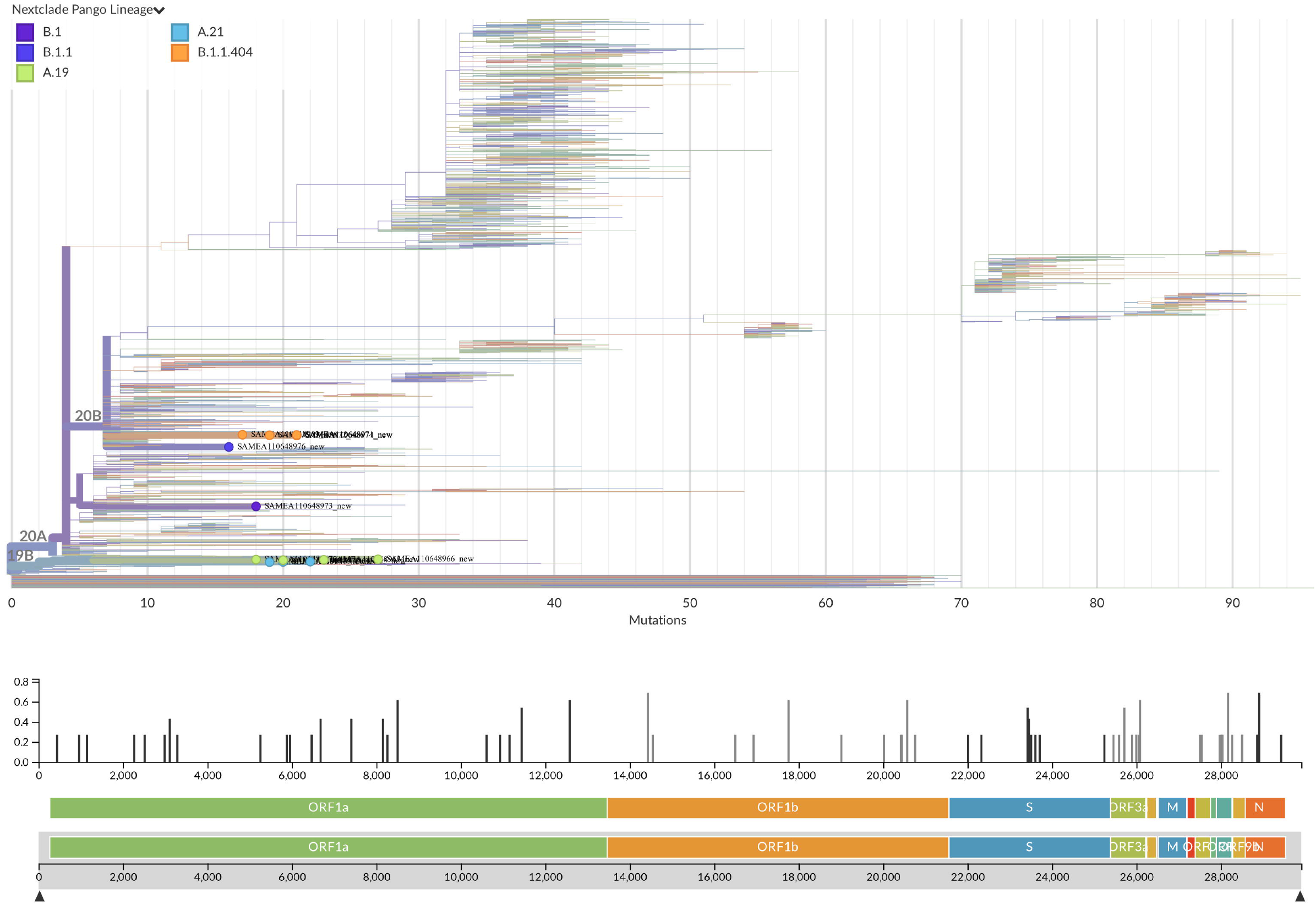
Phylogenetic tree with the 13 new SARS-CoV-2 Burkina Faso genomes placed on a reference tree of 2022 published sequences from all over the world along with the diversity panel below, a bar-chart showing the variation (i.e. mutations) of the 13 sequences in the genome relative to the reference sequence Wuhan-Hu-1/2019 (MN908947). Phylogenetic tree generated with Nextclade online software v2.5.0 (https://clades.nextstrain.org) (accessed on 13 September 2022) and visualized using Auspice v2.37.3 (https://auspice.us/) (accessed on 13 September 2022).

In order to estimate the frequency of SARS-CoV-2/malaria co-infection for the same period and subjects, malaria was screened by microscopy. 219 individuals out of 998 tested positive for malaria (22% CI 19.4–24.5). Like in the case of SARS-CoV-2, there were significant differences in malaria prevalence between age groups (Kruskal-Wallis chi-squared = 132.18, df = 3, p-value < 0.001), but in this case, malaria was mostly detected in children below 12 years old (42.2%; 95% CI 36.1–48.4) (**Figure 1; Table 1**). In addition, a small sex difference was observed with a higher prevalence in male subjects (W = 92508, p-value < 0.05) (**Supplementary Figure 2**). Similar to the temporal trend observed for SARS-CoV-2, malaria cases also showed a significant increase from 5.26% at the beginning of the study to 29.2% at the end of the sampled period (**Supplementary Figure 3**). This could suggest an impact in malaria control due to the pandemic. Indeed, according to official reports, the incidence of malaria in Burkina Faso (cases per 1000 population at risk) increased from 366.1 in 2019 to 389.9 in 2020^12^. Of the 7 SARS-CoV-2 seropositive/PCR positive samples, none of them were co-infected with malaria. Whereas of the 17 seronegative/PCR positive samples, two co-infections with malaria were reported (**Table 1, Figure 1B**). Finally, of the remaining 25 seropositive/PCR negative subjects, 8 were positive for malaria (**Table 1. Figure 1B**). Since the serology test measures total antibodies (IgG/A/M) for SARS-CoV-2, it cannot differentiate recent or past infections, therefore, we cannot rule out whether these data correspond to a co-infection or a sequential infection.

To summarize, we found 2 cases of confirmed SARS-CoV-2 (PCR positive) co-infection with malaria and 8 cases of suspected co-infection (seropositive/PCR negative). Of them, eight were children and teenagers below 14 years and two were adults of >40 years (**Table 1**). There were no significant differences in coinfection frequency between females and males (6 males, 4 females). Only one of the two PCR positive co-infected samples was suitable for sequencing (Ct < 35) and the linage was A.21.

Similar low frequency for SARS-CoV-2 and malaria co-infections found in our study (1%) has been also reported in other studies (34–38) (**Supplementary Table 4**). A systematic review of COVID-19 and malaria coinfection studies has estimated 11% of pooled prevalence (data of 5 studies) with a high degree of heterogeneity (39). However, it is important to note that these studies examined symptomatic patients at the hospital and our study tested stratified asymptomatic people randomly. A recent study reported a higher prevalence of co-infection (12%) with highest prevalence in the age group of 0–20 years (22%) and above 60 years (20%), in this case malaria infection was evaluated in a cohort of hospitalized patients in Uganda with SARS-CoV-2 PCR-confirmed infection (40). As for the symptomatology of co-infections, preliminary data suggest that patients with SARS-CoV-2 and malaria did not seem to have a worst disease outcome but previous malaria exposure seems to be related to less frequency of severe COVID-19 (40). Another study reports that patients co-infected with malaria had significantly faster recovery compared to those not co-infected (38). One limitation of our study to analyze and draw conclusions about clinical outcomes of the co-infections is that all volunteers were asymptomatic. In addition, the low frequency of SARS-CoV-2 and malaria parasite single infections in the population (the small sample size of volunteers that were positive for either disease), reduces the probability of find them together.

It has been suggested that the low incidence and mortality of COVID-19 in malaria endemic regions could be related to cross-immunity and common immunodominant epitopes between malaria and SARS-CoV-2 (40–43). In-silico analysis have identified potential shared targets for immune response between SARS-CoV-2 and *Plasmodium falciparum* proteins which could generate cross-reactivity through HLA and CD8+ T-cell activation (42). Moreover, tuberculosis and malaria prevalence had been significantly associated with reduced COVID-19 mortality (43). Previous studies have also indicated that malaria-induced immunomodulation could be protective against respiratory viruses, reducing pulmonary inflammatory response inflammation (44,45). Alternatively, the higher burden of infectious diseases in sub-Saharan Africa has been suggested to mediate the asymptomatic SARS-CoV-2 infections by induction of immunological tolerance or trained immunity to other infections such as tuberculosis, other human coronaviruses and even Bacillus Calmette-Guérin (BCG) vaccination (19,43). In addition, other studies have reported that bacterial, fungal and viral co-infections with SARS-CoV-2 are also uncommon, however when present, they may cause a worse outcome (46–48). In the case of coinfections with HIV, the prevalence seems higher (26,6%) and HIV subjects showed an increased risk of hospital admission for COVID-19 (49). However, the studies are still preliminary and further research is needed to test these hypotheses. Therefore, additional prospective studies are required to establish a cause-effect relationship between the SARS-CoV-2 disease outcome and co-infection with other circulating pathogens. These should include more patients, control groups, clinical and immunological information, tracing on SARS-CoV-2 severity when co-occurrence with other prevalent infectious diseases.

## 4 Conclusions

To our knowledge, this is the first population-based SARS-CoV-2 prevalence study performed in Burkina-Faso during the first wave of the COVID-19 pandemic. In addition, this is also one of the few studies that examines the co-occurrence of SARS-CoV-2 infection with the malaria parasite in an asymptomatic population.

The findings from this seroprevalence study for SARS-CoV-2 indicate that the prevalence of antibodies against the new coronavirus was around 3% in Burkina Faso at the time of analysis, with large differences in prevalence at local level and between various age classes. This supports the hypothesis that Africa had one of the lowest coronavirus case rates and that would not be due, at least entirely, to a lower testing capacity. Another important finding of our study is the low frequency of co-infection with malaria among the sampled population (1.0%). Of these, 8 out of the 10 cases reported were in children (4) or teenagers (4), and 2 cases in adults of >40 years.

Finally, the whole genome sequencing analysis revealed that the circulating lineages found in Burkina Faso during the first wave of the pandemic were early clades derived from the Wuhan strain. Most of the lineages reported have been previously described in Burkina Faso or the neighbour countries. However, we also identified two less frequent lineages that were probably imported to Burkina-Faso from USA or Europe. We believe that data presented in this study help to fill the gap of the SARS-CoV-2 epidemiological situation in sub-Saharan Africa, where interactions with endemic infectious diseases are common.

## Data Availability

The datasets generated for this study can be found in the European Nucleotide Archive (ENA) database (https://www.ebi.ac.uk/ena/browser/home) with project ID PRJEB55393 and sample IDs SAMEA110648965-SAMEA110648977.

## 6 Author Contributions

Conceptualization was the responsibility of EG-D that also supervised the project with NI. RSY and JBO provided infrastructure and organized fieldwork activities in Burkina Faso. SRY, DMSS, AFY performed data collection. DL-F and MP-M performed RT-qPCR assays. IC, IG-M and MT-P conducted whole genome sequencing and phylogenetic analysis. DL-F and EG-D analysed the data and wrote the manuscript. NI, SRY and IC reviewed the manuscript. All authors read and approved the final manuscript.

## 8 Acknowledgments

We would like to thank all volunteers who participated in this study, as well as the local authorities and communities in Burkina Faso for their support. We also thank the IPBLN, IRSS and IBV core facilities for their support to project activities.

## 10 Supplementary Material

Supplementary figures 1 to 3.

Supplementary tables 1 to 4

Supplementary references

## Footnotes

1 https://covid19.who.int/

2 https://gisaid.org/submission-tracker-global/

3 https://www.who.int/teams/global-malaria-programme/reports/world-malaria-report-2021

4 https://data.worldbank.org/indicator/SH.MLR.INCD.P3?locations=BF, accessed on 27 April 2022

5 https://www.protocols.io/view/ncov-2019-sequencing-protocol-bp2l6n26rgqe/v1?version_warning=no

6 https://gitlab.com/fisabio-ngs/sars-cov2-mapping

7 https://zenodo.org/record/4069557#.X37nuXUzaWg

8 https://clades.nextstrain.org (accessed on 13 September 2022)

9 https://auspice.us/

10 https://cov-lineages.org/lineage_list.html

