## Supplemental material for "Prevalence of SARS-CoV-2 and co-occurrence/co-infection with malaria during the first wave of the pandemic (the Burkina Faso case)"

### *Supplementary Material*

#### 1 Supplementary Figures and Tables

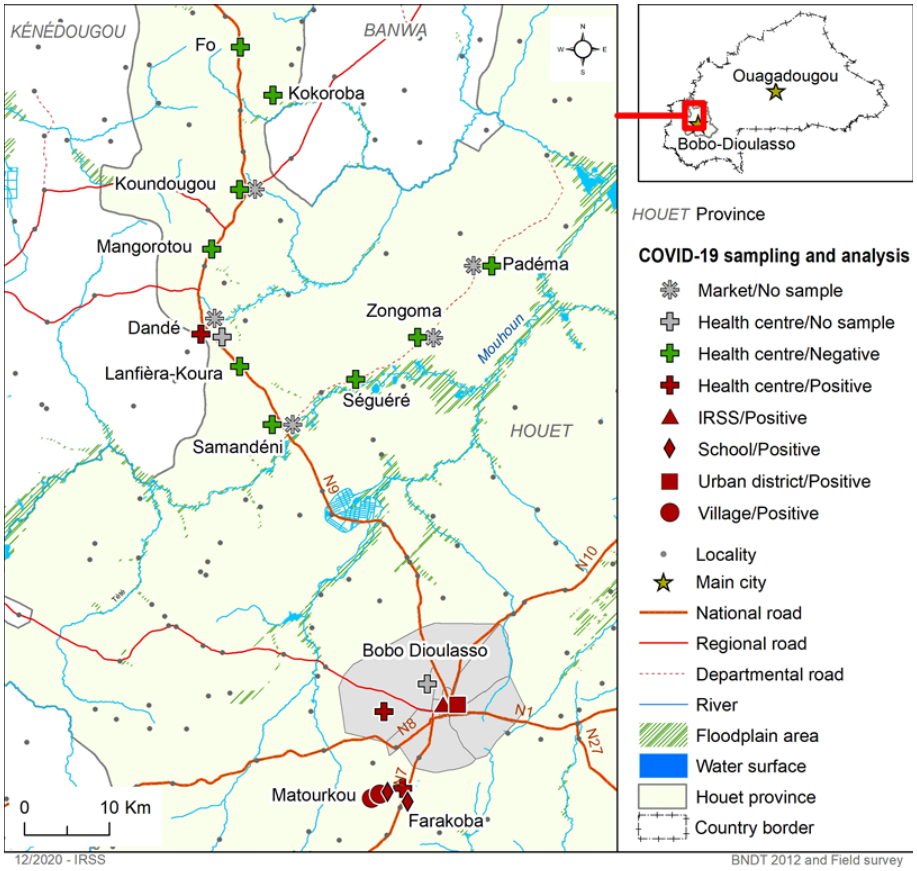


**Supplementary Figure 1.** Map of sampling area in Burkina Faso.

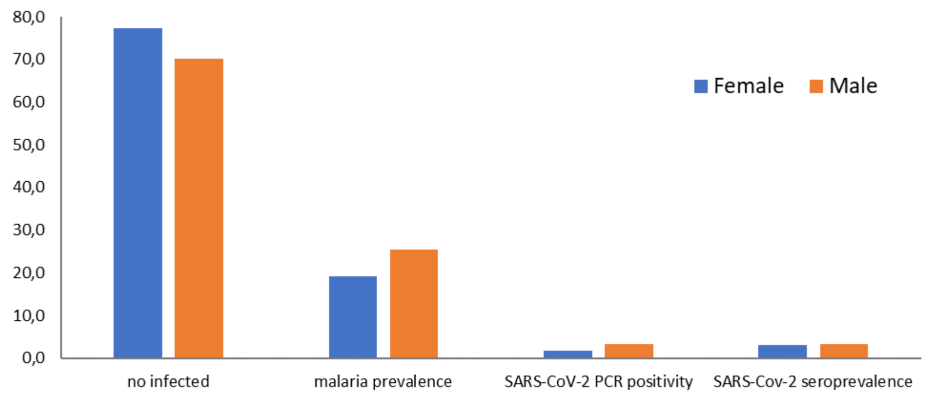


**Supplementary figure 2**. Prevalence of SARS-CoV-2 and malaria positive cases by gender.

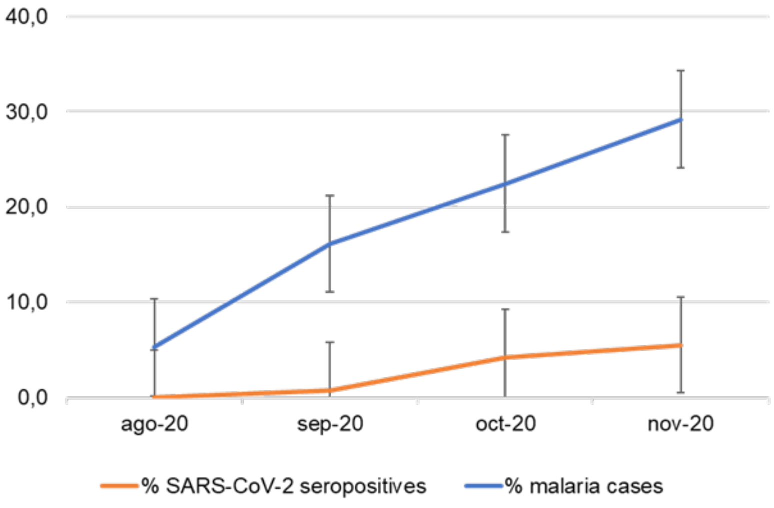


**Supplementary figure 3.** Prevalence of SARS-CoV-2 and malaria over the study period.

**Supplementary table 1.** Representative anti-SARS-CoV-2 seroprevalence studies in Africa. Peer reviewed articles and pre-prints of studies that reported anti-SARS-CoV-2 seroprevalence of general or specific populations in Africa. Reported data of overall seroprevalence. 32 studies.

| Subregion | Study location | Period | Study design | Participants | Sample size | Seroprevalence | Ref. |
|---|---|---|---|---|---|---|---|
| Western Africa | Niger, Nigeria | June 2020 | Cross-sectional | General population | 185 | 25.4% | (1) |
| Western Africa | Calabar, Nigeria | June 2020 | Random | Clinical staff and patients | 66 | 7.6% | (2) |
| Western Africa | Nigeria | August 2020 | Cross sectional non-probability sampling | Blood donors | 113 | 42 % (95% CI: 34–52) | (3) |
| Western Africa | Ibadan, Nigeria | Not indicated | Hospital-based cross-sectional | Health workers | 133 | 45.1% | (4) |
| Western Africa | Enugu, Gombe, Lagos and Nasarawa, Nigeria | October 2020 | Cross-sectional | Households | 10.629 | 25.2% (95% CI 21.8–28.6) Enugu, 9.3% (95% CI 7.0–11.5) Gombe, 23.3% (95% CI 20.5–26.4) Lagos, 18% (95% CI 14.4–21.6) Nasarawa. | (5) |
| Western Africa | Guinea Bissau | November 2020 | Cross-sectional in a cohort. | Health workers | 140 | 18% | (6) |
| Western Africa | Senegal | October-November 2020 | Cross-sectional | Households | 1.463 | 28.4% (95% CI: 26.1-30.8) | (7) |
| Western Africa | Lome, Togo | April-May 2020 | Cross-sectional among high-risk sectors | Healthcare, air transport, police, drivers. | 955 | 0.9% (95% CI: 0.4–1.8) | (8) |
| Western Africa | Ivory Coast | July-October 2020 | Volunteering testing | Gold miners | 1.687 | 25.1% | (9) |
| Western Africa | Sierra Leone | March 2021 | Cross-sectional | Households | 1.893 | 2.6% (95% CI 1.9- 3.4) | (10) |
| Western Africa | Mali | July 2020- January 2021 | Prospective cohort | General population | 2.533 | 58.5% (95% CI: 47.5 to 69.4) | (11) |
| Eastern Africa | Yuba, South Sudan | August-September 2020 | Cross-sectional | Residents | 2.214 | 38.3% (95% CI 31.8–46.5) | (12) |
| Eastern Africa | Addis Ababa, Ethiopia | May 2020 | Convenience | General population | 99 | 3.0%, (95% CI: 0.6–8.6) | (13) |
| Eastern Africa | Dire Dawa, Ethiopia | June-July 2020 | Cross-sectional | Adults | 684 | 3.2% (95 % CI 2.0–4.8) | (14) |
| Eastern Africa | Ethiopia | July 2020 | Cross-sectional | General population | 16.932 | 3.5% (95% CI: 3.2-3.8) | (15) |
| Eastern Africa | Addis Ababa and Jimma, Ethiopia | July-September 2020 | Population-based household | General population | 1.856 | 3.5% (95% CI 1.7-5.4%) Addis Ababa. 1.6% (95%CI 0-4.1%) Jimma. | (16) |
| Eastern Africa | Addis Ababa, Ethiopia | April 2020 | Cross-sectional | General population | 301 | 8.8% (95% CI 5.5-11.6) | (17) |
| Eastern Africa | Kenia | April-June 2020 | National census | Blood donors | 3.098 | 5.6% (95% CI 4.8-6.5) | (18) |

| | | | | | | | |
|---|---|---|---|---|---|---|---|
| Eastern Africa | Nairobi, Kenia | November 2020 | Cross-sectional | Residents | 1.164 | 34.7% (95% CI 31.8-37.6) | (19) |
| Eastern Africa | Blantyre, Malawi | May-June 2020 | Longitudinal | Health workers | 500 | 12.3% (CI 8.2 - 16.5) | (20) |
| Central Africa | Yaounde, Cameroon | October-November 2020 | Cross-sectional | Residents | 971 | 29.2% (95% CI 24.3-34.1) | (21) |
| Central Africa | Gabon | July-October 2020 | Prospective | General population | 1.492 | 36.2% | (22) |
| Central Africa | Brazzaville, Republic of Congo | April-July 2020 | Cross-sectional | General population | 754 | 19.8% | (23) |
| Central Africa | Bukavu, Democratic Republic of Congo | July-August 2020 | Cross-sectional in a cohort. | Healthcare workers | 359 | 41.2% | (24) |
| Northern Africa | Alzintan, Libya | April-May 2020 | Random sampling | Community and healthcare workers | 219 | 2.74% | (25) |
| Northern Africa | Egypt | April-October 2020 | Household cohort | General population | 1.598 | 34.8% | (26) |
| Southern Africa | Zambia | July 2020 | Cross-sectional cluster-survey | Residents | 4.258 | 2.1% (1.1–3.1) | (27) |
| Southern Africa | Zambia | July 2020 | cross-sectional of patients | Outpatients | 1.657 | 8.2% (95% CI 5.1–11.4). | (28) |
| Southern Africa | Cape Town, South Africa | August-September 2020 | Convenience sampling | Workers | 405 | 23.7% | (29) |
| Southern Africa | Eastern Cape, Northern Cape, Free State and KwaZulu Natal, South Africa | January 2021 | Convenience | Blood donors | 4.858 | 62.5% (58.9–66.1) Eastern Cape, 31.8% (25.3-38.3) Northern Cape, 45.5% (39.9-51.1) Free State, 52.1% (49.1-55.2) KwaZulu Natal | (30) |
| Southern Africa | South Africa | November 2020 – April 2021 | Cross-sectional household | HIV-Infected and Uninfected Persons | 7959 | 45.2% (95% CI 43.7- 46.7) | (31) |
| Southern Africa | South Africa | January-May 2021 | Convenience | Blood donors | 16.762 | 47.4% (95% CI 46.2-48.6) | (32) |

**Supplementary table 2.** Detection of samples extracted in pools using the Direct SARS-CoV-2 Realtime PCR Kit (Vircell S.L)

| Sample alias | N gene | E gene | hRNAse |
|---|---|---|---|
| BOB108 | 28.62 | 28.78 | 23.74 |
| BOB108 Pool 1/5 | 31.33 | 31.38 | 26.94 |
| BOB108 Pool 1/10 | 31.86 | 31.92 | 27.94 |
| BOB245 | 34.26 | 34.29 | 25.78 |
| BOB245 Pool 1/5 | 36.56 | 36.99 | 27.45 |
| BOB245 Pool 1/10 | 38.57 | 38.91 | 27.55 |
| Positive control | 29.75 | 28.50 | 28.96 |
| Negative control | NA | NA | NA |

**Supplementary table 3.** Description of lineages found at Burkina Faso in 2020 (https://cov-lineages.org/lineage_list.html)

| Lineage | Most common countries | Earliest date | Description |
|---|---|---|---|
| A.19 | Cote_d'Ivoire 49.0%, Burkina_Faso 40.0%, Italy 3.0%, Australia 2.0%, France 1.0%, | 2020-06-04 | Cote d'Ivoire/ Burkina Faso lineage |
| A.21 | France 26.0%, Burkina_Faso 25.0%, USA 11.0%, Portugal 4.0%, Gabon 4.0% | 2020-04-10 | Mali/ Burkina Faso lineage |
| B.1.1.404 | Burkina_Faso 62.0%, Germany 15.0%, Ghana 6.0%, Italy 6.0%, Togo 3.0% | 2020-05-25 | Ghana, Burkina Faso, Luxembourg, Germany |
| B.1.1.118 | USA 98.0%, Germany 1.0%, Canada 1.0% | 2020-04-10 | US lineage (TX) |
| B.1 | USA 45.0%, Turkey 12.0%, United Kingdom 7.0%, Canada 4.0%, France 3.0% | 2020-01-10 | A large European lineage the origin of which roughly corresponds to the Northern Italian outbreak early in 2020. |

**Supplementary table 4.** SARS-CoV-2 and malaria co-infection published studies

| Ref. | Study location (date) | Study design | Participants (n) | COVID-19 (n) | Malaria coinfection (n) | Malaria Prevalence among COVID-19 | Overall co-infection prevalence | Clinical data |
|---|---|---|---|---|---|---|---|---|
| (33) | Lagos, Nigeria, (April-May 2020) | Cross-sectional study. | Patients suspected with COVID-19 (617) | 121 | 2 | 1.66% | 0.32% | Asymptomatic (52%), mild (48%) |
| (34) | Dutse, Nigeria (March-July 2020) | Cross-sectional study | Participants (74): patients with COVID-19 (54), healthy controls (20) | 54 | 34 | 62.9% | 45.9% | NS |
| (35) | Rivers State, Nigeria (2020) | Cross-sectional study | Patients with COVID-19 (300) | 300 | 300 | 100% | 100% | NS |
| (36) | Kinshasa, Democratic Republic of Congo | Retrospective cohort study | Patients with COVID-19 (160) | 160 | 1 | 0.63% | 0.63% | Mild (57%), moderate (12%), severe (31%) |
| (37) | Uganda (April-October 2020) | Exploratory prospective | Patients with COVID-19 (597) | 597 | 70 | 12% | 12% | Asymptomatic (43%), mild (39%), moderate (8%), severe (8%), critical (3%) |
| (38) | Uganda (March – December 2020) | Prospective Cohort Study | Patients with COVID-19 (270) | 270 | 4 | 1.5% | 1.5% | Symptomatic, mildly symptomatic or asymptomatic |
| (39) | Malawi (April-September 2020) | Prospective Cohort Study | Patients suspected with COVID-19 (87) | 66 (41 PCR+, 25 IgG+/PCR-) | 3 | 4.5% | 3.45% | Severe acute respiratory infection (SARI) |
| (40) | Mumbai, India (April-October 2020) | Retrospective cohort study | Front-line health-care workers (3.711) | 491 | 27 | 5.5% | 0.73% | Mild (73.9%), moderate (12.5%), severe (2.4%) |

NS: not specified

28. Hines JZ, Fwoloshi S, Kampamba D, Barradas DT, Banda D, Zulu JE, et al. SARS-CoV-2 Prevalence among Outpatients during

Community Transmission, Zambia, July 2020 - Volume 27, Number 8—August 2021 - Emerging Infectious Diseases journal - CDC. Emerg Infect Dis [Internet]. 2021 Aug 1 [cited 2022 Jul 6];27(8):2166–8. Available from: https://wwwnc.cdc.gov/eid/article/27/8/21-0502_article

35. Onosakponome EO, Wogu MN. The Role of Sex in Malaria-COVID19 Coinfection and Some Associated Factors in Rivers State, Nigeria. J Parasitol Res. 2020;2020.

36. Matangila JR, Nyembu RK, Telo GM, Ngoy CD, Sakobo TM, Massolo JM, et al. Clinical characteristics of COVID-19 patients hospitalized at Clinique Ngaliema, a public hospital in Kinshasa, in the Democratic Republic of Congo: A retrospective cohort study. PLoS One [Internet]. 2020;15(12 December):1–15. Available from: http://dx.doi.org/10.1371/journal.pone.0244272

37. Achan J, Serwanga A, Wanzira H, Kyagulanyi T, Nuwa A, Magumba G, et al. Current malaria infection, previous malaria exposure,

and clinical profiles and outcomes of COVID-19 in a setting of high malaria transmission: an exploratory cohort study in Uganda. The Lancet Microbe [Internet]. 2022;3(1):e62–71. Available from: http://dx.doi.org/10.1016/S2666-5247(21)00240-8

38. Bakamutumaho B, Cummings MJ, Owor N, Kayiwa J, Namulondo J, Byaruhanga T, et al. Severe COVID-19 in uganda across two epidemic phases: A prospective cohort study. Am J Trop Med Hyg. 2021;105(3):740–4.

39. Morton B, Barnes KG, Anscombe C, Jere K, Matambo P, Mandolo J, et al. Distinct clinical and immunological profiles of patients with evidence of SARS-CoV-2 infection in sub-Saharan Africa. Nat Commun. 2021;12(1).

40. Mahajan NN, Gajbhiye RK, Bahirat S, Lokhande PD, Mathe A, Rathi S, et al. Co-infection of malaria and early clearance of SARS-CoV-2 in healthcare workers. J Med Virol. 2021;93(4):2431–8.
